# Vaccine effectiveness after 1^st^ and 2^nd^ dose of the BNT162b2 mRNA Covid-19 Vaccine in long-term care facility residents and healthcare workers – a Danish cohort study

**DOI:** 10.1101/2021.03.08.21252200

**Authors:** Ida Rask Moustsen-Helms, Hanne-Dorthe Emborg, Jens Nielsen, Katrine Finderup Nielsen, Tyra Grove Krause, Kåre Mølbak, Karina Lauenborg Møller, Ann-Sofie Nicole Berthelsen, Palle Valentiner-Branth

## Abstract

**Background:** At the end of 2020, Denmark launched an immunization program against SARS-CoV-2. The Danish health authorities prioritized persons currently living in long-term care facilities (LTCF residents) and frontline healthcare workers (HCW) as the first receivers of vaccination. Here we present preliminary population based vaccine effectiveness (VE) estimates in these two target groups.

**Methods:** The study was designed as a retrospective registry- and population-based observational cohort study including all LTCF residents and all HWC. The outcome was a polymerase chain reaction confirmed SARS-CoV-2, and VE was estimated for different periods following first and second dose. We used Poisson and Cox regressions to estimate respectively crude and calendar time-adjusted VE for the BNT162b2 mRNA Covid-19 Vaccine from Pfizer/BioNTech with 95% confidence intervals (CI) for vaccinated versus unvaccinated.

**Results:** A total of 39,040 LTCF residents (median age at first dose; 84 years, Interquartile range (IQR): 77-90) and 331,039 HCW (median age at first dose; 47 years, IQR: 36-57) were included. Among LTCF residents, 95.2% and 86.0% received first and second dose from 27 December 2020 until 18 February 2021, for HWC the proportion was 27.8% and 24.4%. During a median follow-up of 53 days, there were 488 and 5,663 confirmed SARS-CoV-2 cases in the unvaccinated groups, whereas there were 57 and 52 in LTCF residents and HCW within the first 7 days after the second dose and 27 and 10 cases beyond seven days of second dose. No protective effect was observed for LTCF residents after first dose. In HCW, VE was 17% (95% CI; 4-28) in the > 14 days after first dose (before second dose). Furthermore, the VE in LTCF residents at day 0-7 of second dose was 52% (95% CI; 27-69) and 46% (95% CI; 28-59) in HCW. Beyond seven days of second dose, VE increased to 64% (95% CI; 14-84) and 90% (95% CI; 82-95) in the two groups, respectively.

**Conclusion:** The results were promising regarding the VE both within and beyond seven days of second vaccination with the BNT162b2 mRNA Covid-19 Vaccine currently used in many countries to help mitigate the global SARS-CoV-2 pandemic.

**Impact of the research:** So far, observational studies of the real-word effectiveness of the mRNA Vaccine BNT162b2 has been limited to the period after the administration of the first dose. This is the first report to date to present vaccine effectiveness (VE) estimates after the second BNT162b2 mRNA Covid-19 Vaccine. We estimated a VE of 52% and 46% in LTCF residents and HCW within seven days, which increased to 64% and 90% in the two groups respectively beyond seven days of immunization. These findings supports maintaining a two-dose schedule of the BNT162b2 mRNA Covid-19 Vaccine.

## Introduction

In Denmark, the national Covid-19 vaccination strategy aims at protecting those at risk of severe outcomes of a SARS-CoV-2 infection resulting in hospitalization or death, and secure key societal functions such as the healthcare system ^1^. Hence, residents in long-term care facilities (LTCF) and front-line healthcare workers (HCW) were target groups prioritized for vaccination, when the vaccination program started on 27 December 2020.

On 16 December 2020 (week 51), a nationwide partial lock down was initiated following widespread community transmission of SARS-CoV-2. In week 53 when the immunization program started, there were several reports of Covid-19 outbreaks in LTCF and the positive percentage for tested LTCF residents was 4.21% (Figure 1a) while it was 2.21% for HCW (Figure 1b).

**Figure 1a and 1b:**
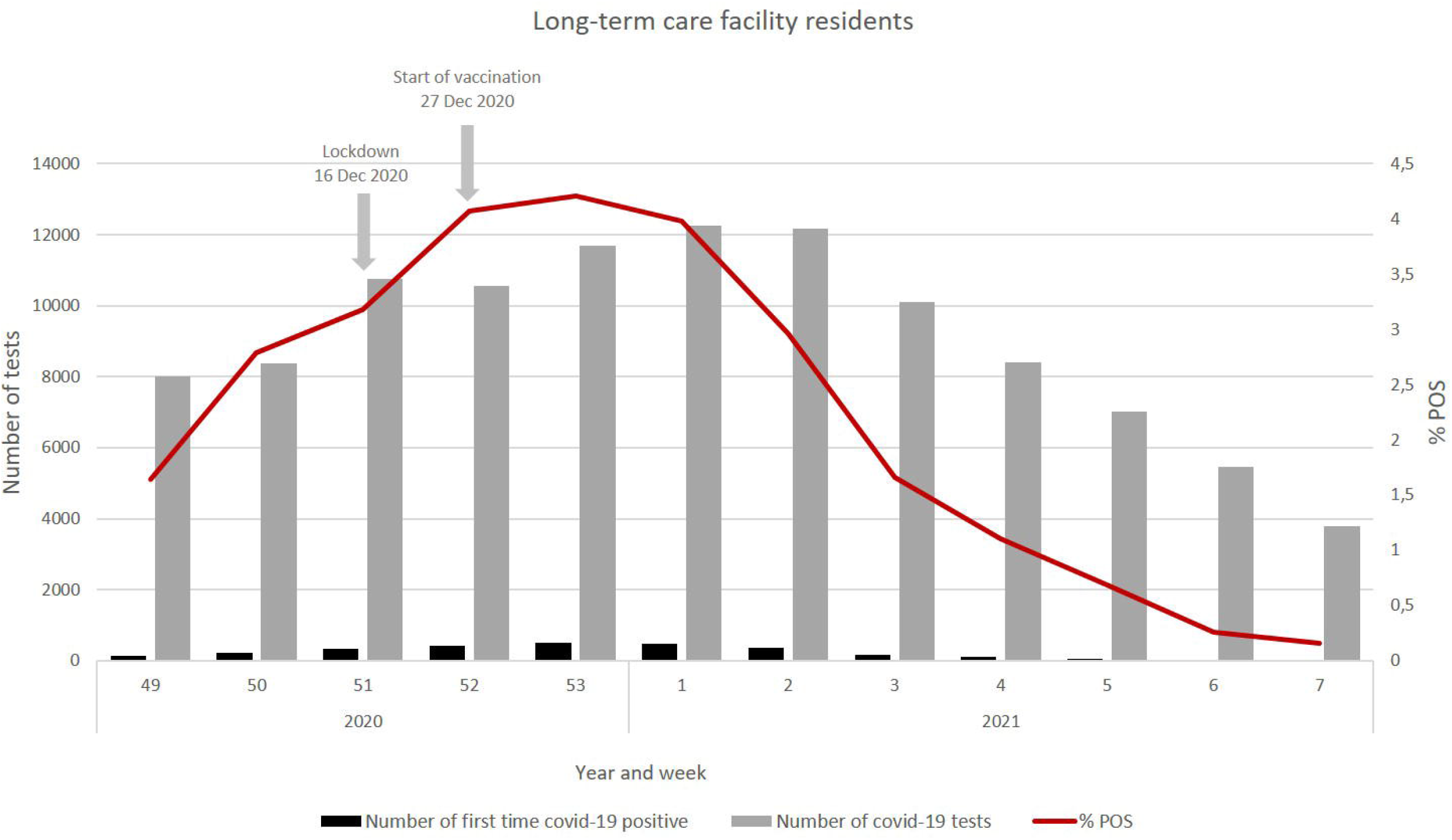
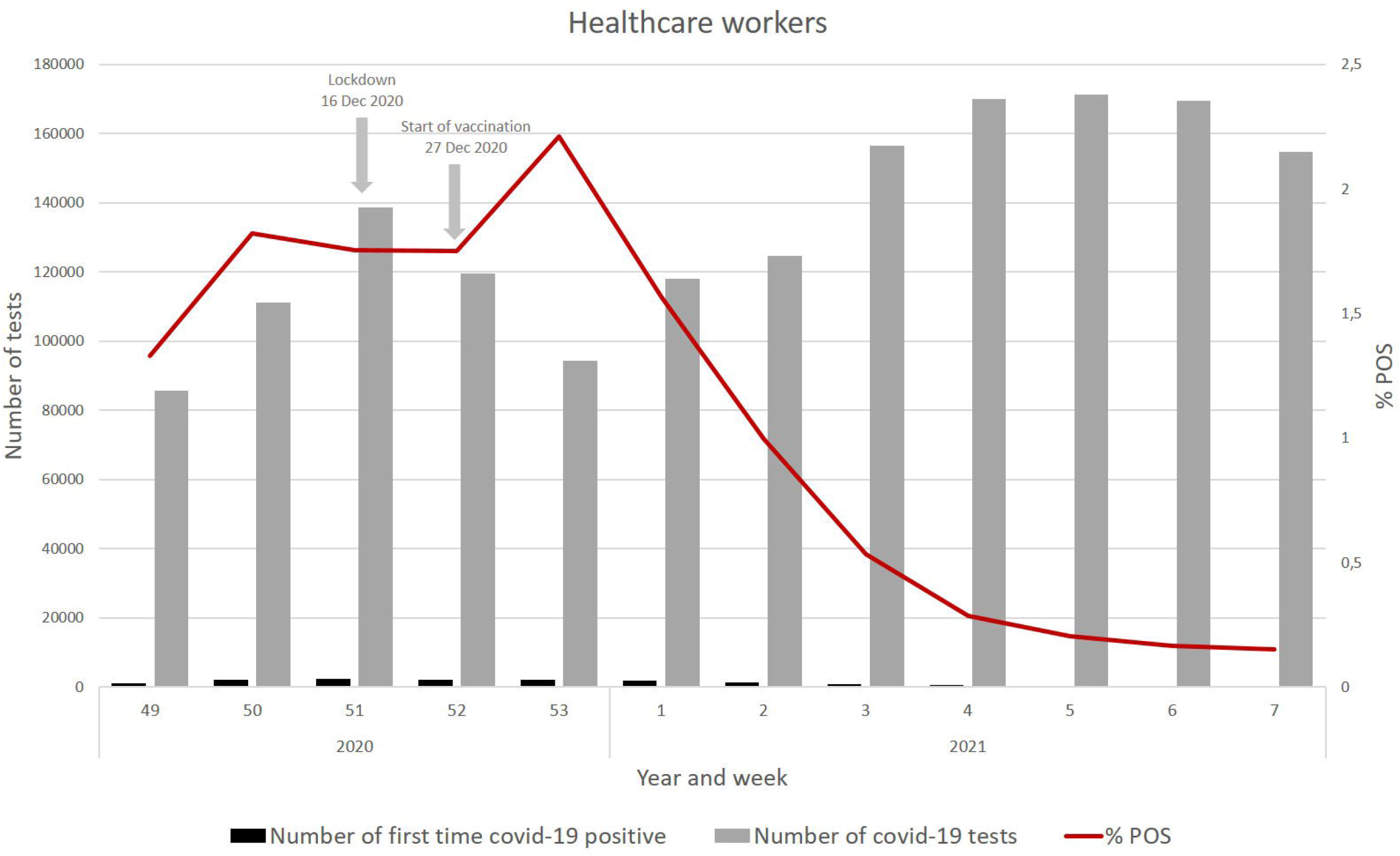
Weekly number of laboratory confirmed SARS-CoV2, number of tests and proportion of positive tests distributed on test weeks for LTCF residents and HCW in Denmark from week 49, 2020 to week 7, 2021. Number of tests include a maximum of one test per day per person until and including the first positive test. Tests of LTCF residents have been identified through The Danish Health and Medicines Authority. Tests of Healthcare workers have been identified through The Danish Agency for Labour Market and Recruitment, in which the current work affiliation is used (updated December 2020).

The aim was to estimate VE for first and second dose of the BNT162b2 mRNA vaccine in two separate cohorts: 1) all LTCF residents, and 2) all HCW, both with no history of SARS-CoV-2, using data from Danish registries.

## Methods

### Data

All individuals living in Denmark are registered in The Danish Civil Registration System with a unique identifier allowing individual-level linkage between various registers ^2^. In this study information on date of birth, immigration and emigration, sex and vital status was collected from The Danish Civil Registration System ^3^. Information on laboratory confirmed SARS-CoV-2 infection was retrieved from the National Danish Microbiology Database (MiBa), where real-time reverse transcription polymerase chain reaction (RT-PCR) test results are registered ^4^. Information on all administered BNT162b2 mRNA vaccines was retrieved from the Danish Vaccination Registry ^5^. Presence of comorbidity within five years (data retrieved in October 2019) was identified based on ICD-10 codes registered for all hospital admissions in The National Patient Registry ^6^. LTCF residents was identified by linking the Civil Registry System to addresses for LTCF by the Danish Health and Medicines Authority ^1 7^. Information on persons working in healthcare was retrieved from The Danish Agency for Labour Market and Recruitment (described in ^8 9^).

### Analyses

We used retrospective population-based cohort studies to estimate VE after first and second vaccine dose. The two cohorts included all LTCF residents and all HCW either living in Denmark at start of vaccination (27 December 2020) or immigrating before the end of study on 18 February 2021. Persons with PCR confirmed SARS-CoV-2 before start of the study were excluded. The cohort was followed from the beginning of the immunization program on 27 December 2020 and until date of positive SARS-CoV-2 test, first dose mRNA-1273 (Moderna) or ChAdOx1 (AstraZeneca) vaccination, emigration, death or end of follow-up (18 February 2021), whichever came first. Receiving first and second dose BNT162b2 mRNA was included in the analysis as time-varying exposures. Summary findings for BNT162b2 mRNA presented by the European Medicines Agency was used to identify relevant time periods for analysis ^10 11^; 1) 0-14 days after first dose (no protection observed) 2) >14 days after first dose and until second dose (partial protection) 3) 0-7 days after second dose (not previously evaluated) 4) > 7 days after second dose (highest protection). The BNT162b2 mRNA vaccine was available from 27 December 2020, the mRNA-1273 (Moderna) vaccine from 14 January 2021 and ChAdOx1 (AstraZeneca) from 9 February 2021. Only BNT162b2 mRNA was included in this study as the other vaccines had too short follow-up. We used a Poisson regression to estimate crude VE and Cox Proportional Hazards regressions to estimate VE adjusted for either age or calendar time.

Data was analyzed using R version 4.0.3 (R Foundation for Statistical Computing, https://www.R-project.org/)

## Results

A total of 39,040 persons (median age at time of first dose: 84 years, IQR (77-90), 63.6% female) were included in the LTCF residents cohort (Table 1). From 27 December 2020 through 18 February 2021, 37,172 LTCF residents received the first dose of the vaccine and 33,567 received the second dose. The HCW cohort included 331,039 persons (median age at time of first dose: 47 years, IQR (36-57), 82.2% female) (Table 1). During the study period, 91,865 HCW received first dose and 80,839 received second dose of the BNT162b2 mRNA vaccine (Table 1). Days between first and second dose for LTCF residents and HCW were 24 (IQR; 20-52) and 25 (IQR 20-51), respectively. More than 99% of the vaccines administered to LTCF residents were BNT162b2 mRNA, which was the case for 89% in HCW (on 18 February 2021).

**Table 1.**
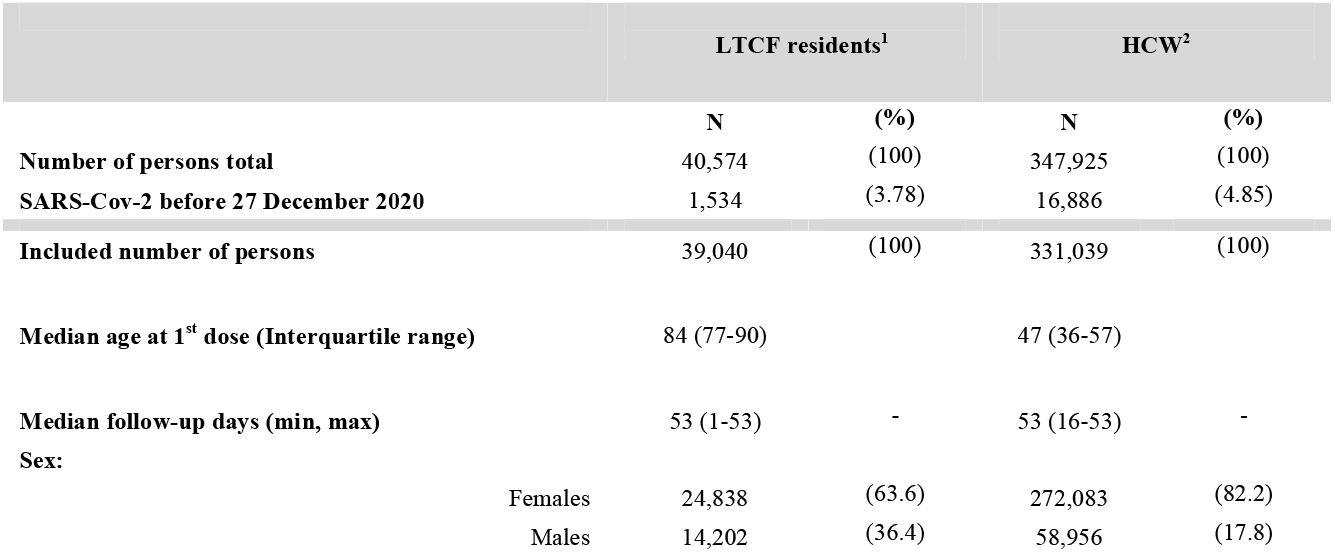
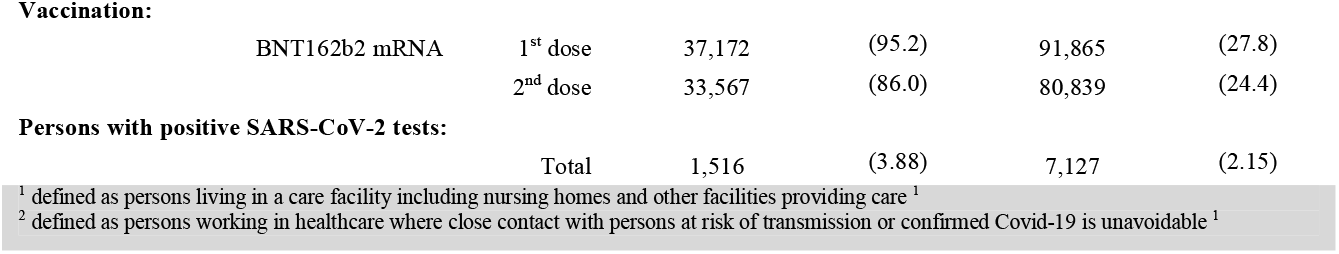
Descriptive overview of median age, median follow-up and SARS-CoV-2 confirmation by vaccination status in 39,040 LTCF residents and 331,039 HCW followed from 27 December 2020, to 18 February 2021 in Denmark.

No significant VE was observed in LTCF residents at any time point between first and second dose after adjusting for calendar time (Table 2). From day 0-7 after second dose, the adjusted VE (VE_adj_) was 52% (95% CI; 27-69) in LTCF residents and after 7 days after second dose it increased to 64% (95% CI; 14-84) for this group (Table 2). A moderate VE_adj_ of 17% (95% CI; 4-28) was observed in HCW from 14 days after first vaccination until second dose (Table 2). VE_adj_ for HCW was 46% (95% CI; 28-59) in day 0-7 after second dose and increased to 90% (95% CI; 82-95) from 7 days after second dose. (Table 2). Adjusting for age, sex and comorbidities did not affect the VE estimates significantly (data not shown). The unadjusted VE was markedly higher than VE adjusted for calendar time, both among LTCF residents and HCW (Table 2).

**Table 2.** Vaccine effectiveness with accompanying 95 % confidence intervals for LTCF residents and HCW after first and second vaccine dose

|  |  |  |  | Unadjusted VE<sup>2</sup> |  | Adjusted VE<sup>3</sup> |  |
| --- | --- | --- | --- | --- | --- | --- | --- |
|  | No. of persons with<br>SARS-CoV-2<sup>1</sup> | PYRS | IR | VE | 95% CI | VE | 95% CI |
| **Nursing home residents** |  |  |  |  |  |  |  |
| Unvaccinated | 488 | 1059.09 | 0.46 | - | - | - | - |
| 0-14 days after 1<sup>st</sup> dose | 760 | 1388.32 | 0.55 | -0.19 | -0.44;0.01 | -0.40 | -0.62;-0.02 |
| > 14 days after 1<sup>st</sup> dose until 2<sup>nd</sup> dose | 184 | 1008.17 | 0.18 | 0.60 | 0.46;0.71 | 0.21 | -0.11;0.44 |
| 0-7 days after 2<sup>nd</sup> dose | 57 | 633.5 | 0.09 | 0.80 | 0.70;0.88 | 0.52 | 0.27;0.69 |
| > 7 days after 2<sup>nd</sup> dose | 27 | 1435.9 | 0.02 | 0.96 | 0.91;0.98 | 0.64 | 0.14;0.84 |
| **Health care workers** |  |  |  |  |  |  |  |
| Unvaccinated | 5663 | 37599.85 | 0.15 | - | - | - | - |
| 0-14 days after 1<sup>st</sup> dose | 1189 | 3425.09 | 0.35 | -1.13 | -1.69;-0.97 | -1.04 | -1.18;-0.91 |
| > 14 days after 1<sup>st</sup> dose until 2<sup>nd</sup> dose | 213 | 2821.84 | 0.08 | 0.50 | 0.29;0.66 | 0.17 | 0.04;0.28 |
| 0-7 days after 2<sup>nd</sup> dose | 52 | 1525.19 | 0.03 | 0.77 | 0.57;0.90 | 0.46 | 0.28;0.59 |
| > 7 days after 2<sup>nd</sup> dose | 10 | 2468.95 | 0.00 | 0.97 | 0.90;100 | 0.90 | 0.82;0.95 |

<sup>1</sup>Laboratory confirmed SARS-CoV-2 test<sup>2</sup>Crude VE estimate<sup>3</sup>VE adjusted for calendar time**Discussion**

## Discussion

For LTCF residents, we found an adjusted VE of 52% (95% CI; 27-69) within 0-7 days after second dose, which increased to 64% (95% CI; 14-84) from 7 days after second dose. In HCW, we observed the same pattern; an adjusted VE of 46% (95% CI; 28-59) in day 0-7 that increased to 90% (95% CI; 82-95) after 7 days following the second dose.

It is very reassuring that we find high VEs in a real-world setting where one cohort had a median age of 84 years at time of first vaccination The findings for the LTCF cohort are encouraging as this group comprises the most vulnerable part of the population and immunosenescence in the elderly population is generally a challenge for vaccines ^12^.

When the vaccination program was initiated, the national Covid-19 incidence was at the highest level since the beginning of the pandemic and the proportion of LTCF residents tested positive for Covid-19 was increasing (Figure 1a). However, it is worth noting that a partial lockdown had been initiated to reduce circulation of Covid-19 a few weeks before the immunization program started. These interventions introduced consecutively might explain the higher VE observed in the unadjusted analysis. The calendar time adjusted analysis prevents bias related to the comparison of unvaccinated Covid-19 cases observed at the start of the vaccination campaign (characterized by high Covid-19) with Covid-19 cases observed later on where the Covid-19 incidence had decreased.

When vaccination started, some LTCF residences were dealing with Covid-19 outbreaks, why vaccination was postponed in these facilities. This selection for vaccination based on Covid-19 outbreaks may potentially inflate the observed VE. The effects of the national lockdown implemented before Christmas became visual in the beginning of January 2021, where positive test rates decreased among LTCF residents and HCW (Figure 1a and 1b). Therefore, the rate of infection decreased during the study period. The high vaccination coverage achieved in LTCF may have decreased risk of infection within the facilities, as they are relatively closed settings.

The main strengths of this study was that the two population-based cohorts of LTCF residents and HCW were followed using national, high quality information on vaccination status and laboratory SARS-CoV-2 positive samples. Due to the situation at nursing homes, there was increased focus on testing in LTCF. Furthermore, there was a national testing strategy, where HCW were offered weekly PCR-testing. Therefore, we believe that our study includes close to all cases of SARS-CoV-2. Further, our cases included both asymptomatic and symptomatic disease based on all conducted laboratory PCR SARS-CoV-2 tests, nationally. Test results from rapid antigen tests and PCR tests at private test centers were not available. However, they constitute a small fraction of test conducted in the study cohorts. Antigen tests were only offered in limited extent to some HCWs.

The adjusted VE for the HCW cohort presented here, is similar to the vaccine efficacy of 95% (95% CI; 90-98) reported by Polack et al. ^10^ in a phase III trial including 43,448 participants with a median age of 52 at time of vaccination. Another interesting finding that aligned with the phase III trial was the unaffected VE estimates after adjustment for age, sex and comorbid conditions, which may be explained by these populations assumingly being relatively young and healthy at time of vaccination ^10^. Expectedly, the estimated VE among LTCF residents was lower (VE; 64%, 95% CI; 14-84) than the VE reported by Polack and colleagues. This could be explained by the higher vulnerability and age distribution in our cohort, a median age of 84 years in LTCF residents compared to 52 years among the trial participants. It is also evident from studies of influenza vaccines, that vaccines are less effective in the elderly ^12^. We observed a moderate, statistically significant, VE for healthcare workers of 16% for the period from 14 days after first dose until second dose. A similar, though higher, VE was found in a study from Israel at day 13-24 after first dose (VE; 51%, 95% CI; 7.2-78). The results from the two studies suggest a possible moderate effect after first dose. Hunter et al. (UK) reanalyzed the data presented by Chodick et al. using a different approach ^13^, and they found an increasing VE from day 14 after first dose, peaking at day 21 at 90%, suggesting a very high level of immunity even after first dose of the vaccine ^13^. Recently, another Israeli study have presented VEs for HCW on day 1-14 after first dose compared to day 15-24 in vaccinated compared to unvaccinated persons. This study observed a VE of 75% in the latter time period and 35% in the first period. The authors note that 55% of the study population had received second dose on day 21-22, but still interpret the findings as related to first dose.

We observed no or low VE after just one dose. However, it is important to note that there was a short interval (median 24 and 25 days) between first and second dose, and therefore it is unlikely that the full impact of a single dose can be extrapolated from our findings. Our findings compared to findings from studies including only first dose corroborate that a two-dose schedule to obtain maximum benefit of the BNT162b2 mRNA vaccine is important. Nonetheless, studies with a longer follow-up between first and second dose are warranted in order to explore potential advantages of a strategy with a postponed second dose.

The cohorts included in this study are two different segments of the Danish population. The findings on VE in LTCF residents may apply to other vulnerable elderly populations with similar conditions. This is the first study to report VE in LTCF residents, an age group not included in previous studies, where the populations were relatively young. The estimated VEs for HCW might be more generalizable as this population in many ways is comparable with other subpopulations. HCW have special knowledge on importance of protective equipment, why the VE in this group may not extrapolate to all populations in the same age groups. On the other hand, HCW have an increased risk of infection, through patient contacts, that may be higher than in the general population.

All in all, the present study confirms, from the field, that two doses of BNT162b2 mRNA vaccine offers protection against SARS-CoV-2 infections in two critical groups, albeit with a better VE among HCW with a median age of 47 years than among a vulnerable population of long-term-care facility residents with a median age of 84 years.

## Data Availability

Data was provided from Danish registries that are not readily available to everyone. Researchers can apply for data through 'Forkermaskinen' at The Danish Health Data Authority.

https://sundhedsdatastyrelsen.dk/da/forskerservice/forskermaskinen

## Acknowledgements

The authors are grateful to The Danish Health Data Authority for their help in defining the population-cohorts. Special thanks to Mette Lange, Line Hansen, and Joanna Phermchai-Nielsen. We would also like to thank the Department of Data Integration and Analysis at Statens Serum Institut for excellent data management.

## References

1. Health TNBo. Retningslinje for håndtering af vaccination mod COVID-19 [red. Guidelines for handling vaccination against Covid-19]: The National Board of Health; 2021 [Available from: https://www.sst.dk/-/media/Udgivelser/2021/Corona/Retningslinjer/Retningslinje-for-haandtering-af-vaccination-mod-COVID.ashx?la=da&hash=9F4DE3E6CB26472FD8977007D4BCF75D925094BC accessed 17/2/2021 2021.

2. Pedersen CB. The Danish Civil Registration System. Scandinavian Journal of Public Health 2011;39(7_suppl):22–25. doi: 10.1177/1403494810387965

3. Authority DHD. The Central Person Registry: The Ministry for Economic Affairs and the Interior; [Available from: https://www.danishhealthdata.com/find-health-data/CPR-registeret accessed 17/2/2021 2021.

4. Voldstedlund M, Haarh M, Molbak K, et al. The Danish Microbiology Database (MiBa) 2010 to 2013. Euro Surveill 2014;19(1) doi: 10.2807/1560-7917.es2014.19.1.20667 [published Online First: 2014/01/18]

5. Grove Krause T, Jakobsen S, Haarh M, et al. The Danish vaccination register. Euro Surveill 2012;17(17) doi: 10.2807/ese.17.17.20155-en [published Online First: 2012/05/04]

6. Lynge E, Sandegaard JL, Rebolj M. The Danish National Patient Register. Scand J Public Health 2011;39(7 Suppl):30–3. doi: 10.1177/1403494811401482 [published Online First: 2011/08/04]

7. Authority TDHaM. Residents in nursing homes [Danish web page] [Available from: https://plejehjemsoversigten.dk/ accessed 14/02/2021 2021.

8. Recruitment TDAfLMa. DREAM vejledning version 44 v2 [red. DREAM guidlines version 44 v2] [Available from: file:///C:/Users/IRMH/Downloads/DREAM%20koder%20-%20%20version%2044%20-%20E%20(3).pdf accessed 17/2/2021 2021.

9. Denmark S. eIndkomstRegister 2021 [Available from: https://www.dst.dk/da/Statistik/dokumentation/Times/eindkomstregister accessed17/2/2021 2021.

10. Polack FP, Thomas SJ, Kitchin N, et al. Safety and Efficacy of the BNT162b2 mRNA Covid-19 Vaccine. N Engl J Med 2020;383(27):2603–15. doi: 10.1056/NEJMoa2034577 [published Online First: 2020/12/11]

11. Agency EM. Assessment report Comirnaty Common name: COVID-19 mRNA vaccine (nucleoside-modified) 2020 [Available from: https://www.ema.europa.eu/en/documents/assessment-report/comirnaty-epar-public-assessment-report_en.pdf accessed 22/02/2021 2021.

12. Osterholm MT, Kelley NS, Sommer A, et al. Efficacy and effectiveness of influenza vaccines: a systematic review and meta-analysis. Lancet Infect Dis 2012;12(1):36–44. doi: 10.1016/s1473-3099(11)70295-x [published Online First: 2011/10/29]

13. Hunter PR, Brainard J. Estimating the effectiveness of the Pfizer COVID-19 BNT162b2 vaccine after a single dose. A reanalysis of a study of ‘real-world’ vaccination outcomes from Israel. medRxiv 2021:2021.02.01.21250957. doi: 10.1101/2021.02.01.21250957

